# Genomic Medicine Guidance: A Point-of-Care App for Heritable Thoracic Aortic Diseases

**DOI:** 10.1101/2023.12.22.23299696

**Authors:** Rohan Patil, Fatima Ashraf, Samer Abu Dayeh, Siddharth K. Prakash

**Affiliations:** Department of Internal Medicine, McGovern Medical School, University of Texas Health Science Center at Houston, Houston, Texas, USA; McWilliams School of Bioinformatics, University of Texas Health Science Center at Houston, Houston, Texas, USA

**Author notes:** **Correspondence:** Siddharth K. Prakash, MD, PhD, McGovern Medical School, University of Texas Health Science Center at Houston, 6431 Fannin Street, MSB 6.116, Houston, Texas 77030. These authors contributed equally to this work (Rohan Patil, Fatima Ashraf).

**Keywords:** genomic medicine, point-of-care, thoracic aortic aneurysm, aortic dissection, decision support

## Abstract

Genetic testing can determine familial and personal risks for heritable thoracic aortic aneurysms and dissections (TAD). The 2022 ACC/AHA guidelines for TAD recommend management decisions based on the specific gene mutation. However, many clinicians lack sufficient comfort or insight to integrate genetic information into clinical practice. We therefore developed the Genomic Medicine Guidance (GMG) app, an interactive point-of care tool to inform clinicians and patients about TAD diagnosis, treatment, and surveillance. GMG is a REDCap-based app that combines publicly available genetic data and clinical recommendations based on the TAD guidelines into one translational education tool. TAD genetic information in GMG was sourced from the Montalcino Aortic Consortium, a worldwide collaboration of TAD centers of excellence, and the NIH genetic repositories ClinVar and ClinGen. The app streamlines data on the 13 most frequently mutated TAD genes with 2,286 unique pathogenic mutations that cause TAD so that users receive comprehensive recommendations for diagnostic testing, imaging, surveillance, medical therapy, preventative surgical repair, as well as guidance for exercise safety and management during pregnancy. The app output can be displayed in a clinician view or exported as an informative pamphlet in a patient-friendly format. The overall goal of the GMG app is to make genomic medicine more accessible to clinicians and patients, while serving as a unifying platform for research. We anticipate that these features will be catalysts for collaborative projects that aim to understand the spectrum of genetic variants that contribute to TAD.

## 1 Introduction

The global incidence of thoracic aortic dissections, 3 to 6 cases per 100,000 person-years, is almost certainly underestimated because up to half of individuals who experience a dissection die before reaching a hospital (1). In 2022, 7% of out-of-hospital deaths were caused by acute aortic dissections. In more than 20% of cases, a single genetic mutation that is frequently inherited can cause thoracic aortic aneurysms or dissections (TAD, 2). Genetic testing for TAD can lead to earlier recognition of patients who are at risk for acute aortic dissections and timely individualized interventions that can prevent deaths. Technological advances have made genetic testing more practical for clinical use (3, 4). Gene-based management of TAD was codified in the recent ACC/AHA guidelines for aortic disease, which provide different recommendations for diagnosis, surveillance, and preventative aortic surgery depending on the mutated gene (5).

However, there are numerous barriers to implementing genetic information into routine clinical workflows (2). Genetic counseling services are not readily accessible to clinicians or patients. Many clinicians are not prepared to integrate genetic test results into clinical decision making. In a recent survey, more than 75% of cardiovascular genetic tests were ordered by providers who do not have formal training in clinical genetics (6). Due to these gaps in care, patients frequently do not receive appropriate pre-or post-test counseling to understand test results or adhere to guideline-based recommendations (7).

Point of care tools synthesize complex medical information to guide clinical decision making (8). In genomic medicine, point of care guidance can lead to more frequent and effective utilization of genetic information in clinical practice (9, 10, 11). To address genomic medicine care gaps related to TAD, we created Genomic Medicine Guidance (GMG), a point-of-care application that consolidates and clarifies evidence-based recommendations for clinical management of TAD based on genetic test results. Users who select a pathogenic mutation receive a comprehensive report that describes associated clinical features and recommendations for diagnosis, surveillance, medical therapies, and interventions. We illustrate how the features of the GMG app can lower barriers to integration of genomic concepts into clinical practice and improve the recognition and treatment of TAD.

## 2 Materials and Methods

### 2.1 Project Design

The GMG app was implemented in a version of Research Electronic Data Capture (REDCap) that is hosted at the McWilliams School of Biomedical Informatics at UTHealth Houston. REDCap is a secure, web-based application designed to support data capture for research studies, providing 1) an intuitive interface for validated data entry; 2) audit trails for tracking data manipulation and export procedures; 3) automated export procedures for seamless data downloads to common statistical packages; 4) procedures for importing data from external sources; and 5) a MySQL database server to facilitate data queries and retrieval. Curated variant information and clinical recommendations are stored in the MySQL database. The user interface is implemented in PHP, a robust and widely used scripting language. The database can be updated continuously by users who enter new variant data through the interface or by project managers who can upload bulk variant data using the data import tools in REDCap.

### 2.2 App Code

The GMG app is a custom PHP plugin that interacts with the REDCap framework to display, extract, add to, or modify the SQL database. The plugin sequentially queries the SQL database for gene and variant information or associated clinical content in response to user queries. The GMG plugin is rendered in HTML, JavaScript (JS), and CSS. In particular, the app makes use of jQuery, a JS library. When a user directly interfaces with the JS selection fields, AJAX methods in jQuery are leveraged to dynamically load data in the background and update the display without reloading the entire web page. AJAX methods make use of built-in PHP developer functions in REDCap to query the database and return the appropriate information.

The tooltip class is used to display additional information about predefined terms such as acronyms when the user hovers the mouse pointer over text. Results can be formatted for viewing on different screens (phone, desktop, iPad) the Bootstrap v5.1.2 framework for CSS. We created download and print functions by customizing a PHP script that retrieves data from the SQL database and converts the data to PDF format using the dompdf package. The results file can be emailed to users using built-in REDCap functions.

## 3 Results

### 3.1 GMG Data Structure

GMG data fields consist of the gene name, variant information, evidence for pathogenicity, associated clinical features, gene-specific diagnostic workup, variant-specific diagnostic workup, gene-specific surveillance, variant-specific surveillance, gene-specific medical therapy recommendations, gene-specific aortic intervention thresholds, guidance during pregnancy, and activity recommendations. New or revised data can be imported in a batch format using the REDCap import tool (Supplemental Data).

### 3.2 GMG Genetic Content

The GMG app currently includes data on 2,270 pathogenic and likely pathogenic variants in 13 TAD genes: ACTA2, PRKG1, TGFBR1, TGFBR2, SMAD3, MYLK, TGFB2, MYH11, FOXE3, LOX, MFAP5, COL3A1, and FBN1 (Table 1).

### 3.3 User Workflow

Users may freely access GMG at https://go.uth.edu/GMG. To facilitate point of care queries, the home page provides a prompt to select one HTAD gene and one variant from curated dropdown lists (Figure 1). Users may view sample test report forms from commercial genetic laboratories with highlighted variant information. After variant selection, users are required to verify the clinical features that prompted the genetic test and to attest that appropriate pre-test genetic counseling was provided. If users enter a variant that is not on the curated list, the app interface provides an option for them to upload the variant data and associated phenotypic information. The output tab describes the selected variant, including evidence for pathogenicity in ClinVar, clinical features associated with the variant, including lifetime risks for aortic surgery or dissection, and evidence-based clinical recommendations for diagnostic and surveillance imaging of the aorta, medical therapy, and size thresholds for elective aortic surgery. The recommendations are based on the 2022 ACC/AHA Aortic Disease guidelines. The app displays clinician and patient-friendly outputs in different browser tabs that can be printed, downloaded, or emailed in PDF format.

**Figure 1.**
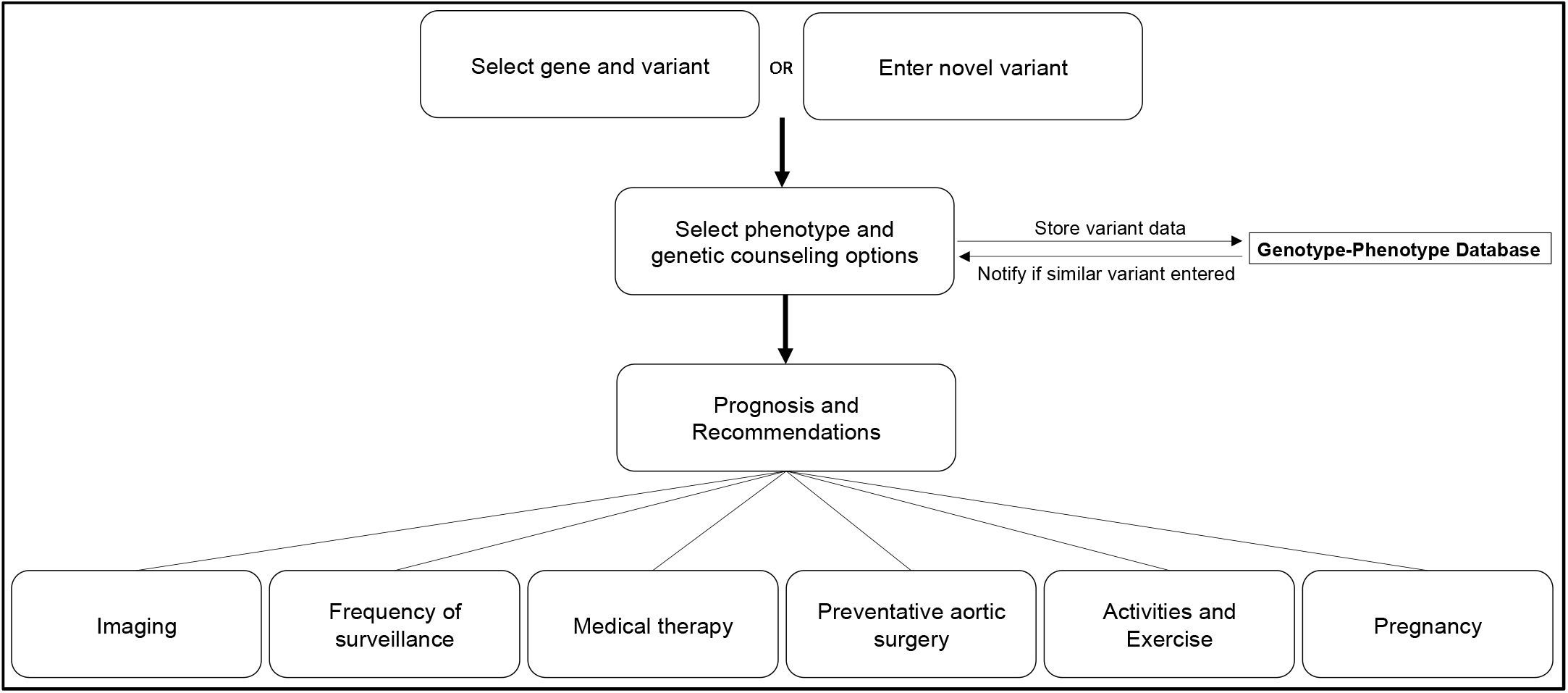
GMG app workflows. Users may select from a list of curated variants or enter a new variant in one of 13 TAD genes. The app is available at https://go.uth.edu/GMG.

### 3.4 Researcher Workflow

Researchers can enter new data into the GMG app or update current app data using REDCap data import tools. Batch genetic and clinical data for multiple gene variants can be uploaded simultaneously into the GMG genotype-phenotype database using a single data import template spreadsheet (Supplemental Data).

### 3.5 Genetic Counseling Content

GMG app users are required to verify that patients were appropriately selected for genetic testing and received counseling about the benefits, risks, and implications of genetic testing. Users can also access genetic counseling resources from within the app, including guidance to locate a genetic counselor for telehealth consultation. Additional links to the Marfan Foundation and the Montalcino Aortic Consortium, two advocacy groups with resources for newly diagnosed patients with TAD, are provided on the output page.

### 3.6 User Reviews

Thirty-eight GMG users completed a Qualtrics feedback survey about their experiences with the app. Users uniformly rated four components of the GMG app as very easy to use: the graphical interface (85/100), selection of a curated gene variant (87/100), understanding clinical recommendations (86/100), and entering a new clinical variant (89/100). Users also rated the clinical recommendations as extremely helpful for clinical guidance (90/100) and highly likely to influence their clinical practice (86/100). Users wrote that some genetic and clinical data in the app may be challenging for individuals who do not have formal training in genetics. They also submitted design recommendations to increase user engagement. Other suggestions included outreach to clinics and professional organizations to establish a user network and an instructional video to orient new users.

## 4 Discussion

Thoracic aortic aneurysms predispose to deadly aortic dissections without obvious physical signs or symptoms and are frequently inherited as single gene mutations. Genetic testing for TAD can identify affected relatives and save lives but is underutilized because genetic services are not available to many families who are at risk. In this report, we describe a new point-of-care app, GMG, that is intended to expand access to genetic information about genetic cardiovascular diseases including TAD. As reflected by user ratings, GMG simplifies the interpretation of genetic data for clinicians who do not have genetic expertise and provides valued clinical guidance based on the test result.

Several recent studies and a review of 20 clinical decision support tools for genomic medicine concluded that they improve risk assessment and lead to meaningful changes in clinical decisions (9,11). Examples of such tools include an app that synthesizes data on somatic mutations in tumors to target cancer therapies based on gene-specific prognosis (10). The Electronic Medical Records and Genomics (eMERGE) network integrates contextual decision support into the electronic medical record to streamline access to genetic testing (12).

GMG includes a genotype-phenotype database that will also promote collaborations by connecting providers who enter similar genetic variants to resolve variants of uncertain significance or build case series to elucidate new disease phenotypes. Crowdsourcing through GMG will expand the clinical and genetic content over time. Future versions of GMG will incorporate gene-based care guidance for other adult-onset genetic cardiovascular diseases that are primarily managed by non-expert clinicians, such as hyperlipidemias, cardiomyopathies, and channelopathies.

## 5 Conclusions

The GMG app expands access to genetic information in an era when most cardiovascular genetic tests are not supervised by genetic professionals. GMG will prevent deaths due to genetic cardiovascular diseases by promoting evidence-based guidelines for gene-based decision making and familial testing. The app also empowers patients to take an informed role in healthcare decisions.

## Data Availability

All data produced in the present study are available upon reasonable request to the authors.

## 6 Conflict of Interest

The authors declare that the research was conducted in the absence of any commercial or financial relationships that could be construed as a potential conflict of interest.

## 7 Author Contributions

RP: Data curation, Writing – Original Draft; SA: Data curation; FA: Resources, Software, Writing – Review & Editing; SP: Conceptualization, Supervision, Funding acquisition, Writing – Review & Editing.

## 8 Funding

This work was supported in part by UL1TR003167.

## 9 Acknowledgments

The authors are grateful to Dr. Elmer Bernstam for project oversight and support.

## Notes

### Competing Interest Statement

The authors have declared no competing interest.

