## Supplementary figures and images for "Genomic Medicine Guidance: A Point-of-Care App for Heritable Thoracic Aortic Diseases"

### Supplemental Table 1

#### 1 Supplementary Material

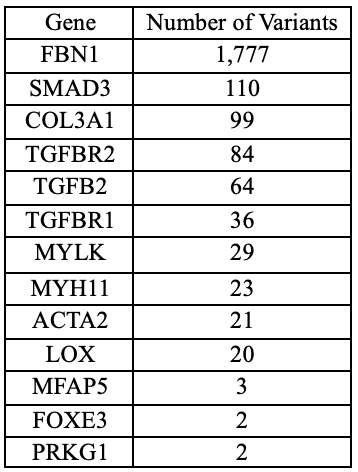

Table 1. Curated variants in the GMG app
